# Profiling an integrated network of cellular senescence and immune resilience measures in natural aging: a prospective multi-cohort study

**DOI:** 10.1101/2023.08.25.23294589

**Authors:** Natalia Mitin, Amy Entwistle, Anne Knecht, Susan L. Strum, Allison Ross, Kirsten Nyrop, Hyman B. Muss, Denis Tsygankov, Joseph M. Raffaele

## Abstract

**Background:** Biological aging begins decades before the onset of age-related clinical conditions and is mediated by both cellular senescence and declining adaptive immune function. These processes are functionally related with the rate of senescent cell accumulation dependent upon a balance between induction and immune clearance. We previously showed that biomarkers in these domains can identify patients at-risk of surgery-related adverse events. Here, we describe evidence of clinical relevance in early aging and metabolic phenotypes in a general adult population.

**Methods:** We enrolled a total of 482 participants (ages 25-90) into two prospective, cross-sectional healthy aging cohorts. Expression of biomarkers of adaptive immune function and cellular senescence (SapereX) was measured in CD3+ T cells isolated from peripheral blood.

**Findings:** We established a network of biomarkers of adaptive immune function that correlate with cellular senescence and associate with early aging phenotypes. SapereX immune components associated with a decrease in CD4+ T cells, an increase in cytotoxic CD8+ T cells, and a loss of CD8+ naïve T cells (Pearson correlation 0.3-0.6). These components also associated with a metric of immune resilience, an ability to withstand antigen challenge and inflammation. In contrast, SapereX components were only weakly associated with GlycanAge (Pearson correlation 0.03-0.15) and commonly used DNA methylation clocks (Pearson correlation 0-0.25). Finally, SapereX biomarkers, in particular p16, were associated with chronic inflammation and metabolic dysregulation.

**Interpretation:** Measurement of SapereX biomarkers may capture essential elements of the relationship between cellular senescence and dysregulated adaptive immune function and may provide a benchmark for clinically relevant health decisions.

## Introduction

Aging is a complex process that begins decades before the onset of common age-related conditions such as frailty and overt chronic disease^1–3^. As such, many aspects of aging remain poorly understood. However, the immune system is a central mediator of organismal aging with well-described early changes beginning in the second or third decade. Therefore, biomarkers of immune system aging are needed to identify early vulnerabilities and allow targeted interventions to both repair immune system function and improve an individual’s aging trajectory.

Immune aging is comprised of immunosenescence, characterized by changes in the proportion and function of numerous immune cell types, as well as replicative or cellular senescence. Cellular senescence is an umbrella term describing a cellular response to a wide variety of stimuli including genomic instability, telomere attrition, epigenetic changes, and impaired proteostasis^4^. Cells in this state are characterized by stable growth arrest, resistance to apoptosis, and a complex, pro-inflammatory secretome. Cellular senescence promotes aging, and the removal of senescent cells reverses aging pathology^5–7^. Seminal work by Yousefzadeh and Niedernhofer showed that induction of cellular senescence solely in immune cells can induce cellular senescence in diverse cell and tissue types throughout the body, impairing their function^8^.

Cells that have undergone cellular senescence accumulate with age. The rate of accumulation depends on both the rate of induction by the stressors described above, as well as the capacity of the immune system, particularly cytotoxic CD8+ T cells, to surveil and clear these altered cells^9,10^. In biologically young, healthy organisms, cellular senescence is maintained in an adaptive balance. As the organism ages or the immune system is impaired due to other challenges, cellular senescence accumulates and causes dysfunction. While multiple lines of evidence from independent labs suggest a role for the adaptive immune system in the maintenance of cellular senescence, the relationship is complex and is just beginning to be understood.

Previous studies showed little association between cellular senescence and traditional measures of immunosenescence^1,11^. We took a different approach, interrogating a curated list of biomarkers known to reflect adaptive immune function and compared them to the expression of p16, the most uniquely associated biomarker of cellular senescence, as well as longitudinal changes in p16 (ref^12^, Justice et al, in print; Sapere Bio, unpublished). Biomarkers that correlated with p16 individually were retained for further analysis as potential biomarkers of immunosurveillance of senescent cells. In the first evidence of clinical relevance, the resulting network of biomarkers was used to identify patients at risk for adverse events associated with cardiac surgery^12^. Here, we continue to interrogate and characterize this set of biomarkers to establish a broader relationship between cellular senescence and adaptive immune system function. We now describe evidence of clinical relevance in early aging and metabolic phenotypes in a cohort of healthy adults.

## Materials and Methods

### Study design and participants

Two cohorts were enrolled. Community-dwelling adults were enrolled into a natural aging registry study (community cohort). Patients seeking physician-guided health interventions for longevity were enrolled into a long-term registry study (physician-guided cohort). Adults between the ages of 25-85 years old living in North Carolina were recruited into the community cohort. Exclusion criteria were diagnosis of an autoimmune disorder, history or current or chemotherapy, immunotherapy, or radiation therapy, history of solid organ or bone marrow transplants, dialysis, or an active infection (acute or chronic) for which antibiotics and/or antivirals were prescribed within the last 14 days. To ensure representation across age ranges among the 250 participants, we enrolled 30 participants in the age group of 25-34 years, 45 in 35-44, 50 in 45-55, 50 in 55-64, 45 in 65-74, and 30 in 75-85. The clinical study was registered with clinicaltrials.gov (NCT05123859). The IRB of the University of North Carolina at Chapel Hill gave ethical approval for this study. Adult patients (25 years and older) seeking care at the Raffaele Medical Clinic were enrolled into a physician-guided cohort. There were no exclusion criteria for medical reasons. An independent IRB (Advarra) gave ethical approval for this study.

### Sample collection and biomarker measurements

Peripheral blood samples were collected at a lab facility (Any Lab Test Now, Durham, NC) or at the Raffaele Medical clinic using Sapere Bio sample collection kits that stabilize whole blood. 7.5ml of blood was used to isolate T cells as previously reported^13^. T cell pellets were stored at -80°C until further analysis. Gene expression of p14, p16, LAG3, CD28, and CD244 was analyzed by real-time qPCR and normalized to housekeeping genes. Positive and negative controls were included in each run, and Cts over 37 were considered below the limit of detection. Expression of each SapereX gene is reported as log2 of arbitrary units, as is standard for qPCR reporting.

### Statistical analyses

Descriptive statistics were reported as mean (standard deviation) for continuous variables and as frequency (%) for categorical variables. For all analyses, missing data for any reason was not imputed. If a measure was missing, the subject was excluded from summary statistics and analyses. Pairwise associations were quantified using the Pearson correlation coefficient and p-value (two-tailed). One-way ANOVA analyses were used where three or more groups were present. Two-tailed p-values of less than 0.05 were considered statistically significant. Statistical analyses were performed in SAS version 9.4 and JMP 12.2.0 (SAS Institute, Cary, NC).

## Results

### Study cohorts

In this study, a total of 482 participants were enrolled in one of two registries – 250 community-dwelling adults (>25 years) were enrolled in a community cohort and 232 patients seeking physician-guided health interventions for longevity were enrolled in a physician-guided cohort. As shown in Table 1, demographics and clinical characteristics were similar across cohorts with some exceptions. The community cohort had more female participants, higher BMI, and a higher rate of diabetes. The physician-guided cohort had a higher rate of hyperlipidemia and pre-diabetes as well as small statistically (but not clinically) significant differences in other measures such as blood pressure, heart rate, hemoglobin, and hematocrit. Notably, the physician-guided cohort had a higher neutrophil-to-lymphocyte ratio, a marker of systemic chronic inflammation^14–16^.

**Table 1.** Participant characteristics.

|  | Community cohort (n=250) | Physician-guided cohort (n=232) | p value |
| --- | --- | --- | --- |
| Age<br>mean+/- sd, years<br>range | 54 +/- 16<br>25 - 85 | 59 +/- 12<br>22 - 94 | <0.0001 |
| Female gender, % | 181 (72.7) | 116 (52.0) | <0.0001 |
| BMI, mean+/- sd, kg/m<sup>2</sup> | 26.0 +/- 5.3 | 24.4 +/- 4.7 | 0.005 |
| Hypertension*, n (%) | 37 (14.9) | 20 (20.0) | 0.34 |
| Syst BP mean+/- sd, mmHg | 119 +/- 18 | 125 +/- 17 | 0.06 |
| Diastolic BP mean+/- sd, mmHg | 78 +/- 10 | 72 +/- 9 | <0.0001 |
| Heart rate mean+/- sd, bpm | 67 +/- 11 | 64 +/- 11 | 0.01 |
| Hyperlipidemia**, n (%) | 97 (38.8) | 94 (51.1) | 0.01 |
| Total cholesterol, mean+/- sd, mg/dL | 192 +/- 36 | 202 +/- 42 | 0.05 |
| HDL-c mean+/- sd, mg/dL | 66 +/- 15 | 68 +/- 20 | 0.8 |
| LDL-c mean+/- sd, mg/dL | 107 +/- 32 | 117 +/- 37 | 0.004 |
| Triglycerides mean+/- sd, mg/dL | 94 +/- 49 | 91 +/- 46 | 1.0 |
| Diabetes, n (%)<br>Diabetes<br>Pre-diabetes<br>No diabetes | 7 (2.8)<br>6 (2.4)<br>237 (94.8) | 2 (2.0)<br>8 (8.2)<br>88 (89.8) | 0.03 |
| HbA1c, mean+/- sd, % | 5.3 +/- 0.5 | 5.4 +/- 0.4 | 0.002 |
| Glucose fasting, mean+/- sd, mg/dL | 91 +/- 14 | 88 +/- 12 | 0.43 |
| HOMA-IR, mean+/- sd, |  | 1.5 +/- 1.0 |  |
| CKD, n (%) | 9 (2.1) | 8 (1.9) | 0.8 |
| Serum creatinine, mean+/- sd, mg/dL | 0.8 +/- 0.2 | 0.9 +/- 0.2 | <0.0001 |
| ALT, mean+/- sd, U/L | 18 +/- 10 | 23 +/- 11 | 0.0002 |
| AST, mean+/- sd, U/L | 20 +/- 6 | 26 +/- 9 | <0.0001 |
| Hemoglobin, mean+/- sd, g/dL | 14.0 +/- 1.3 | 14.5 +/- 1.6 | 0.0002 |
| Hematocrit, mean+/- sd, % | 42.0 +/- 3.5 | 43.1 +/- 4.3 | 0.004 |
| RBC, mean+/- sd, K/mm<sup>3</sup> | 4.6 +/- 0.4 | 4.7 +/- 0.5 | 0.28 |
| WBC, mean+/- sd, K/mm<sup>3</sup> | 5.5 +/- 1.6 | 5.7 +/- 1.8 | 0.17 |
| Neutrophil-to-lymphocyte ratio (NLR), mean+/- sd | 1.9 +/- 0.7 | 2.3 +/- 1.5 | 0.001 |

\* >140 mmHg, \*\* total cholesterol >200 md/dL

**Table 2.**
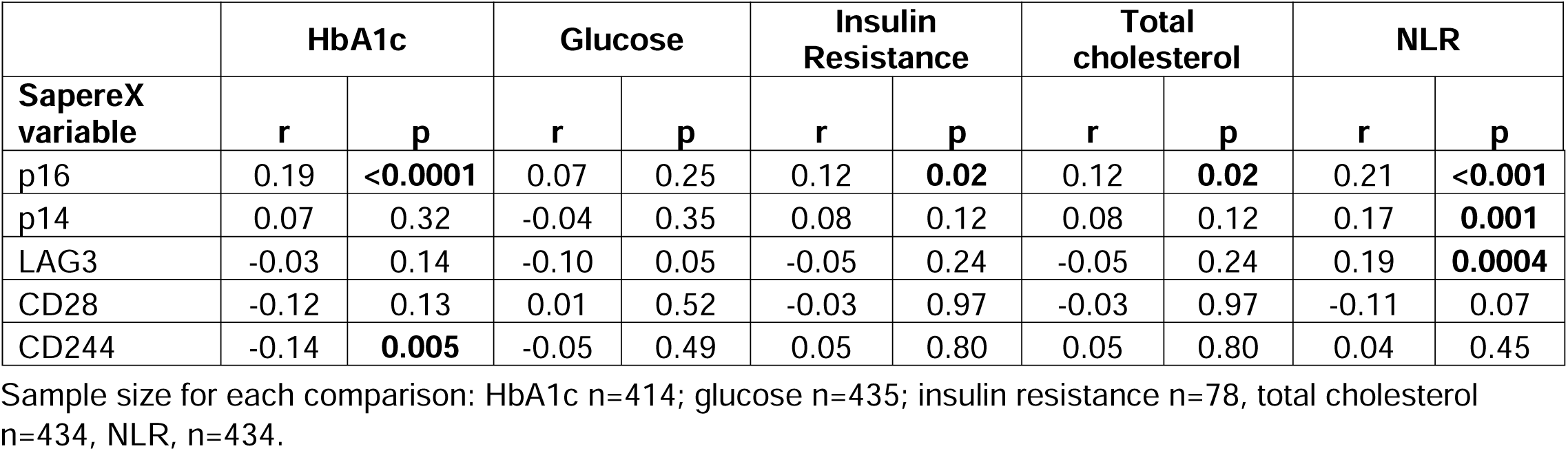
Association between SapereX biomarkers and early pathology.

|  | HbA1c |  | Glucose |  | Insulin Resistance |  | Total cholesterol |  | NLR |  |
| --- | --- | --- | --- | --- | --- | --- | --- | --- | --- | --- |
| SapereX variable | r | p | r | p | r | p | r | p | r | p |
| p16 | 0.19 | **<0.0001** | 0.07 | 0.25 | 0.12 | **0.02** | 0.12 | **0.02** | 0.21 | **<0.001** |
| p14 | 0.07 | 0.32 | -0.04 | 0.35 | 0.08 | 0.12 | 0.08 | 0.12 | 0.17 | **0.001** |
| LAG3 | -0.03 | 0.14 | -0.10 | 0.05 | -0.05 | 0.24 | -0.05 | 0.24 | 0.19 | **0.0004** |
| CD28 | -0.12 | 0.13 | 0.01 | 0.52 | -0.03 | 0.97 | -0.03 | 0.97 | -0.11 | 0.07 |
| CD244 | -0.14 | **0.005** | -0.05 | 0.49 | 0.05 | 0.80 | 0.05 | 0.80 | 0.04 | 0.45 |

Sample size for each comparison: HbA1c n=414; glucose n=435; insulin resistance n=78, total cholesterol n=434, NLR, n=434.

### Relationship between biomarkers of cellular senescence and immune function

We have previously identified a network of biomarkers that link cellular senescence, immune system function, predisposition to surgery-related adverse events^12^. This network included gene expression of p16, p14, LAG3, CD28, and CD244 measured in peripheral blood T lymphocytes. We refer to this network of biomarkers as SapereX. p16 is the biomarker most uniquely associated with cellular senescence and has been used as a marker of established senescent cells in studies linking cellular senescence, functional decline, and aging-related diseases^17,18^. A decrease in CD28^19^ and an increase in LAG3 and CD244 have all been implicated in the loss of immune function, particularly in CD8+ T cells. p14, a transcript related to p16^20^, is a regulator of cell cycle arrest and apoptosis through p53/p21. The pairwise association between biomarkers within SapereX network is shown in Figure 1. Overall, there was a large degree of association between all the components, with CD28 negatively correlating with the other markers. Interestingly, while p16 and p14 were associated with chronological age, other markers were weakly associated and had highly heterogeneous levels of expression in individuals within every age group. Therefore, there is a significant association between levels of a biomarker of cellular senescence (p16) and biomarkers of adaptive immune function.

**Figure 1.**
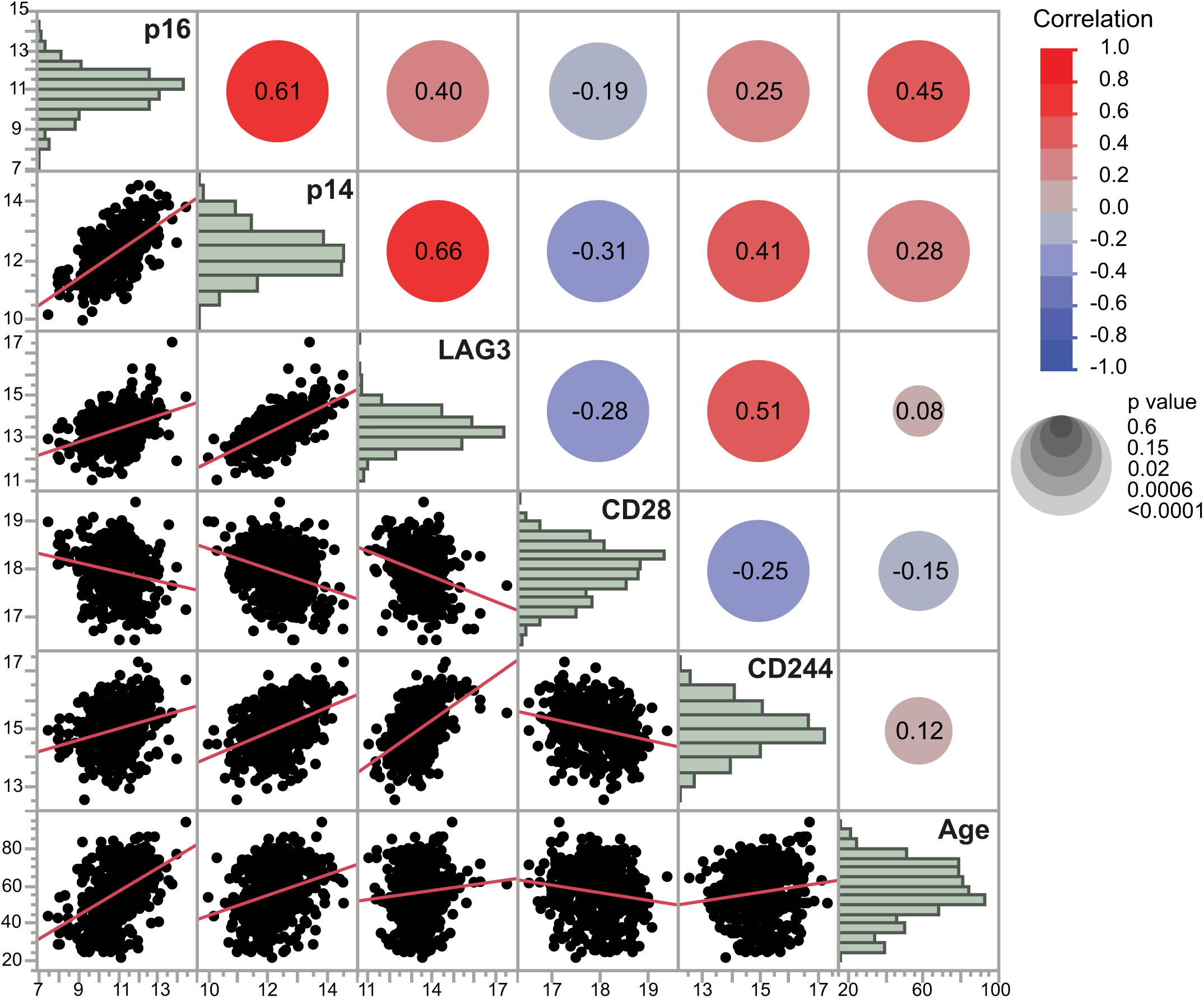
Relationship between biomarkers within SapereX network. Scatterplot correlation matrix of expression levels (log2) of p16, p14, LAG3, CD28, CD244 as well as chronological age. The color of each correlation circle represents the correlation between each pair of variables on a scale from red (+1) to blue (-1). The size of each circle represents the significance test between the variables. A larger circle indicates a more significant relationship, and the Pearson correlation coefficient is shown as a number and a line of linear fit on the corresponding scatterplot. The histograms (diagonal across the matrix) show the distribution of each biomarker in the entire cohort.

### SapereX biomarker network and age-associated immune decline

We also examined the relationship between SapereX and conventional biomarkers of immune function. Traditionally, a decline in immune system function is associated with a decrease in lymphocytes with the corresponding increase in neutrophils, a decrease in CD4+ and an increase in CD8+ cytotoxic T cells, and more specifically an increase of CD8+CD28-T cells that have impaired proliferation and a decrease in CD8+CD95+ naïve T cells. CD4+, CD8+ T cell subset count data via flow cytometry was available in 168 participants in a physician-guided cohort and is shown in Figure 2. As expected, LAG3, CD244, and interestingly, p14 were associated with a decrease in CD4+ and an increase in CD8+ T cells, a decrease in CD8+CD95+ T cells and an increase in CD8+CD28-T cells. CD28 gene expression as measured within SapereX (presence of CD28) was inversely associated with all of the above. Expression levels of p16 were also positively associated with biomarkers of immune decline (a decrease in CD8+CD95+ T cells and an increase in CD8+CD28-T cells) (Figure 2) and also a decline in lymphocytes and an increase in neutrophils.

**Figure 2.**
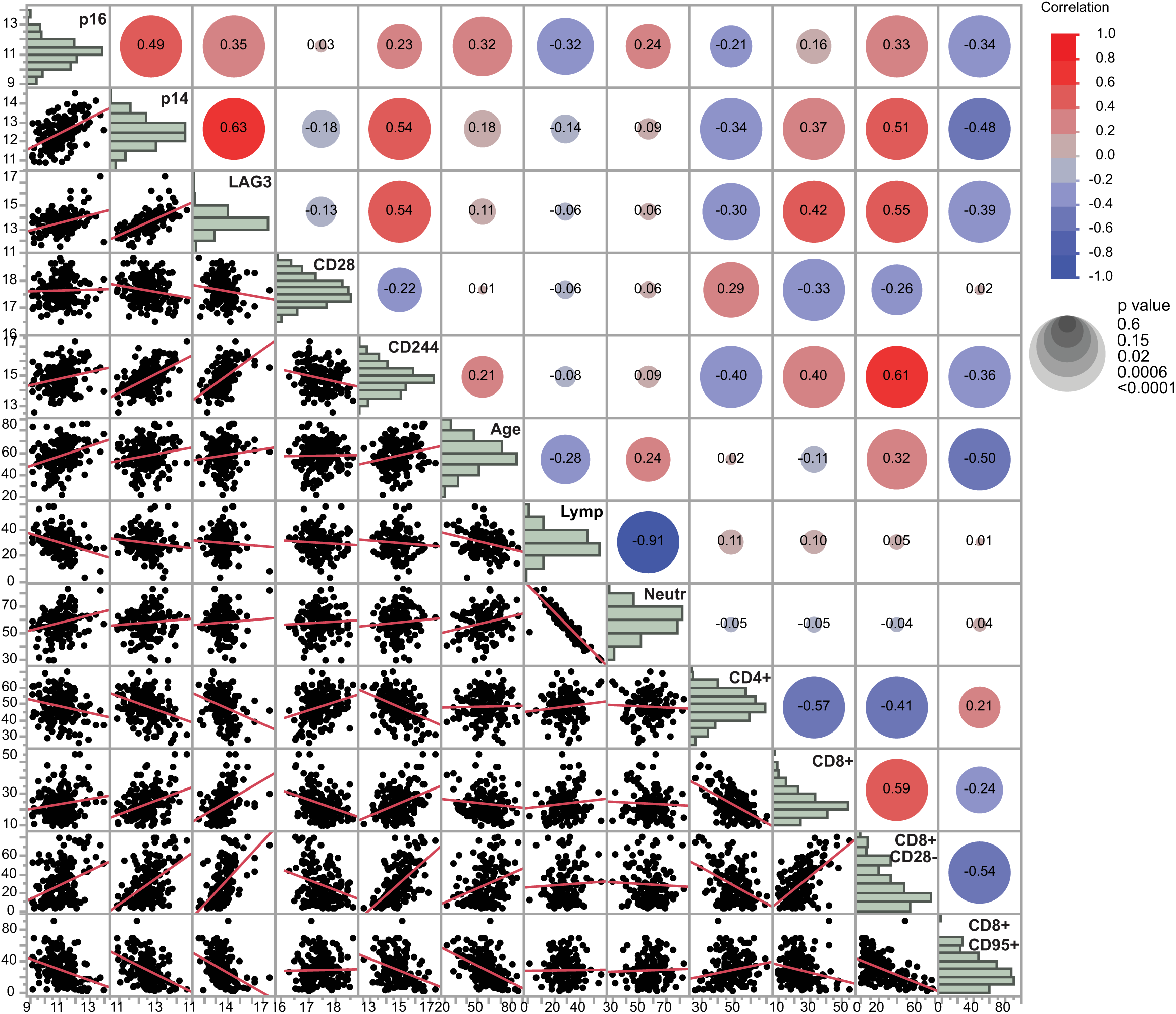
Relationship between SapereX biomarker network and established molecular metrics of age-associated immune decline. Percent of lymphocytes or neutrophils (from CBC labs) as well as CD4+, CD8+, CD8+CD28- and CD8+CD95+ T cells (measured by flow cytometry) were compared to expression levels of SapereX biomarkers. Scatterplot correlation matrix of expression levels (log2) of p16, p14, LAG3, CD28, CD244 as well as chronological age. The color of each correlation circle represents the correlation between each pair of variables on a scale from red (+1) to blue (-1). The size of each circle represents the significance test between the variables. A larger circle indicates a more significant relationship, and the Pearson correlation coefficient is shown as a number and a line of linear fit on the corresponding scatterplot. The histograms (diagonal across the matrix) show the distribution of each biomarker in the entire cohort.

### SapereX and other molecular aging biomarkers

Next, we examined the relationship between SapereX and other biomarkers of aging such as telomere length, intrinsic and extrinsic methylation clocks and GlycanAge. GlycanAge was particularly interesting given its association with immune aging and chronic inflammation. Telomere length was determined as the difference between granulocyte and lymphocyte telomere length measurements to account for individual differences in telomere length (ref). Intrinsic epigenetic aging clock (Fig.3 “Intrinsic”) represents changes in DNA methylation that best fit chronological age. Extrinsic epigenetic clocks (“Extrinsic”, Pace of Aging (PoA) and PhenoAge) are postulated to provide additional information on the aging process. Consistent with our previously published data, there was a weak association between p16 and epigenetic clocks, except for PoA (ref). p16 expression was also weakly positively associated with telomere shortening. There was no association between p16 and GlycanAge. p14 expression showed a similar pattern to p16. There was even less association between the remaining components of SapereX network and other aging biomarkers, except for CD244 and telomere shortening (r=0.34). While, GlycanAge showed no association with any component of the SapereX network, it was associated with the extrinsic epigenetic age clock (TruDiagnostic) and the PhenoAge biological age calculation. Overall, our data suggest that SapereX biomarkers capture different aspects of the aging process than epigenetic clocks, GlycanAge, and to some extent, telomere shortening.

**Figure 3.**
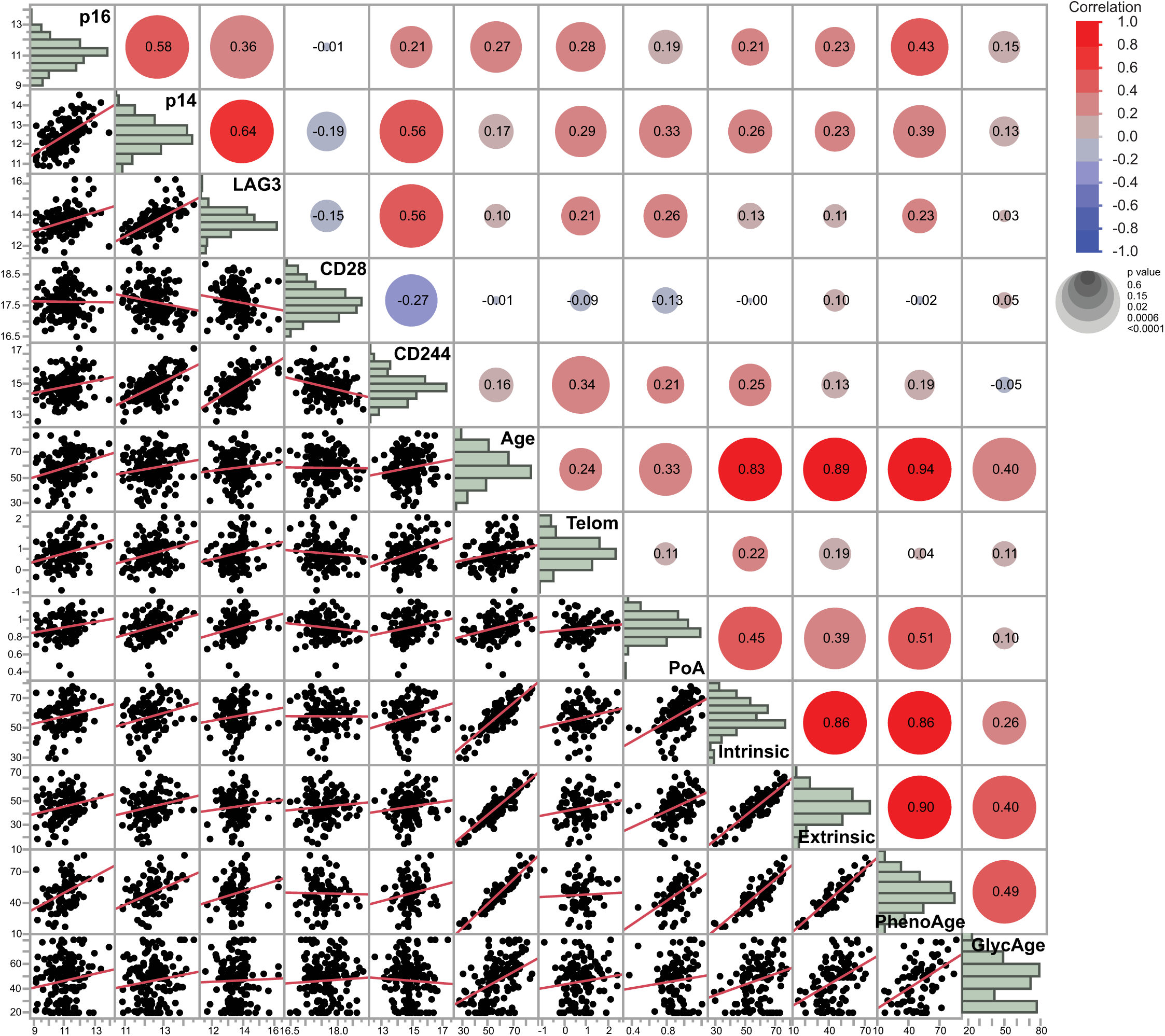
Relationship between SapereX biomarker network and molecular biomarkers of aging. Telomere length, GlycanAge or epigenetic clocks (Pace of aging, intrinsic, extrinsic, PhenoAge) were compared to expression levels of SapereX biomarkers. Scatterplot correlation matrix of expression levels (log2) of p16, p14, LAG3, CD28, CD244 as well as chronological age. The color of each correlation circle represents the correlation between each pair of variables on a scale from red (+1) to blue (-1). The size of each circle represents the significance test between the variables. A larger circle indicates a more significant relationship, and the Pearson correlation coefficient is shown as a number and a line of linear fit on the corresponding scatterplot. The histograms (diagonal across the matrix) show the distribution of each biomarker in the entire cohort.

### SapereX is associated with immune resilience

A recent study of ∼ 48,500 participants developed a quantitative measure of immune resilience linked to longevity and defined as the ability of the immune system to withstand antigen challenges and regulate inflammation^21^. That study weighted CD4+ and CD8+ absolute T cell counts to derive progressively worsening immune grades, with grade I defined as a primordial state that is optimal for immune health. We utilized the same algorithm to categorize immune resilience grades for 168 participants with known CD4+ and CD8+ T cell counts (same subset as Figure 2). Then, we examined the distribution of SapereX biomarkers across grades of immune resilience. LAG3, p14 and CD244 expression progressively increased with the loss of immune resilience (Figure 4). CD28 expression progressively decreased. On the other hand, p16 and chronological age were not independently associated with changes in immune resilience. In support of that idea, PCA analysis (Figure 5) grouped p16 and chronological age in a separate cluster from the rest of the SapereX network.

**Figure 4.**
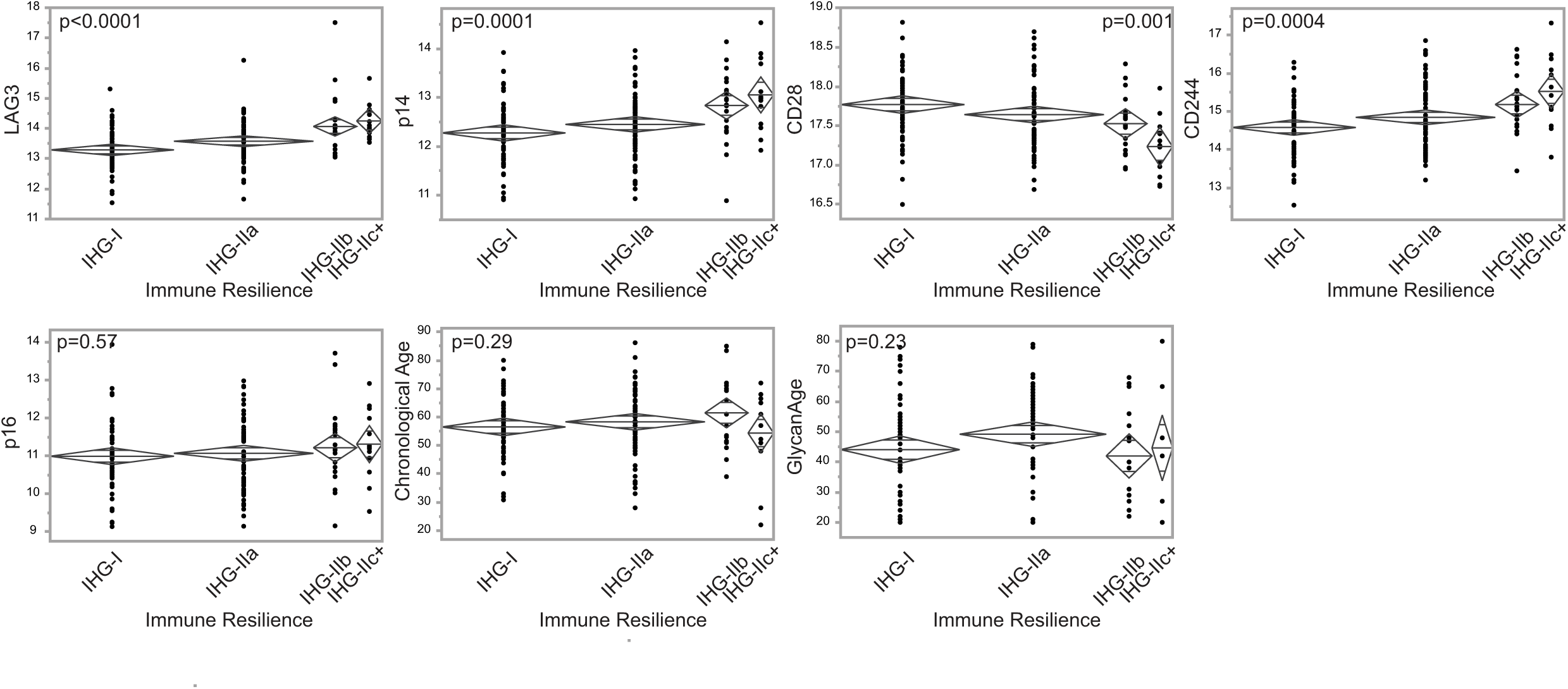
A subset of the SapereX biomarker network is associated with immune resilience. Distribution of SapereX biomarkers, chronological age, or GlycanAge across categories of immune resilience. Immune grades (IHG) were determined based on CD4+ and CD8+ T cell counts and represent grade I, IIa, IIb, and IIc^21^. Participants in Grade I have the most resilience towards antigen-specific challenges and aging in general. p value for one-way ANOVA analysis among groups is shown on each plot.

**Figure 5.**
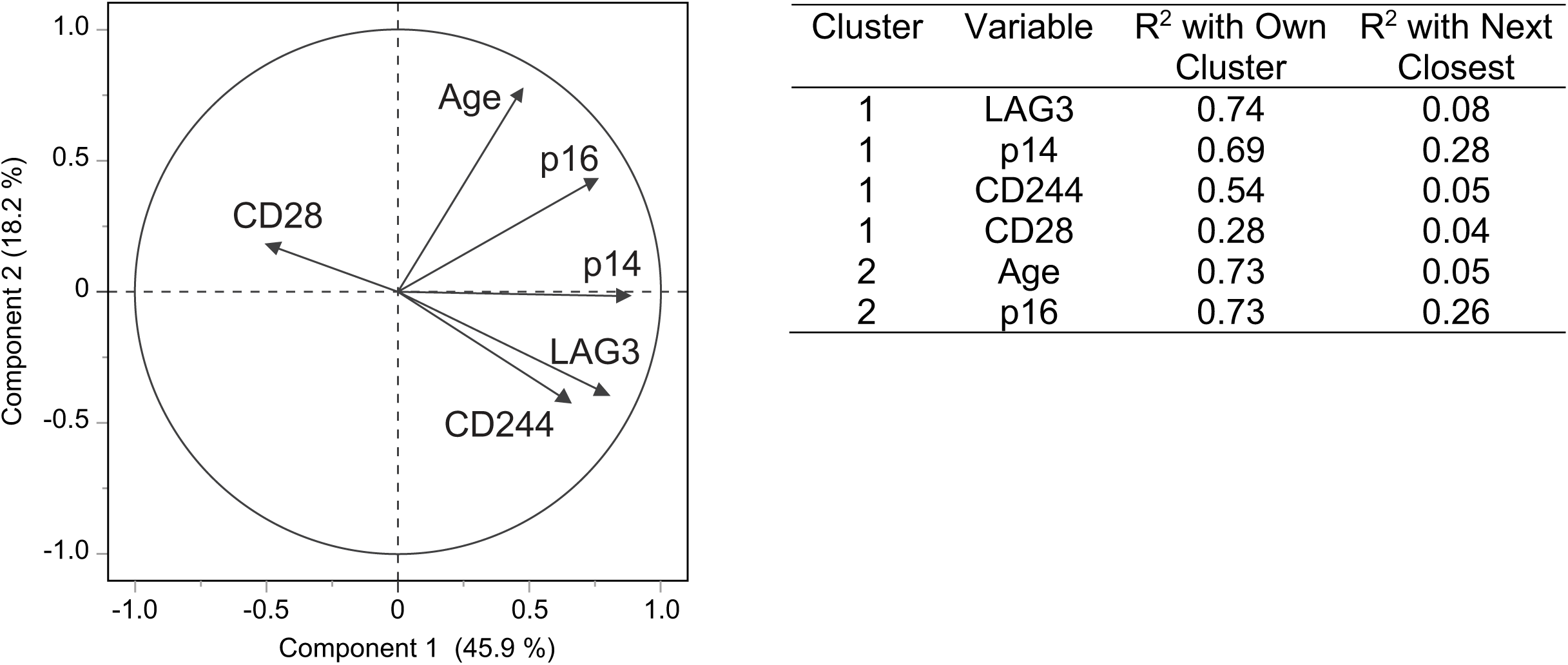
Principal components analysis of SapereX biomarkers and chronological age identified two clusters. P16 and age in cluster #1 and biomarkers associated with adaptive immune system decline and p14 in cluster #2.

Surprisingly, GlycanAge and an extrinsic methylation clock of immune aging (TruDiagnostic) showed no association with immune resilience (Figure 4).

### SapereX is associated with early clinical indicators of decline

We have established that the SapereX network is associated with a number of age-related changes in the adaptive immune system including a new measure of immune resilience. As previously mentioned, a decline in immune system function is linked to the earliest stages of organismal decline, well before the presentation of chronic diseases. We have examined the association between SapereX and clinical markers of physiologic decline. Given the lab results available in these cohorts, we chose biomarkers of metabolic health (insulin resistance, fasting glucose, HbA1C), cardiovascular health (total cholesterol), and neutrophil-to-lymphocyte ratio (NLR), a marker of low-grade systemic inflammation. p16, and not other SapereX components was associated with metabolic health (HbA1C and insulin resistance) and total cholesterol. p16, p14, LAG3, and to a lesser extent CD28 were associated with NLR. None of the SapereX biomarkers were associated with fasting glucose. Association of p16 with insulin resistance is consistent with data in mice where depletion of senescent cells improved insulin sensitivity^22^.

## Discussion

Aging research has historically focused on the very old and on diseases of old age. Recently, attention has shifted to the idea that biological aging begins early and, as such, investigations in subjects across a wide age range and outside the context of reactive disease care are foundational to geroscience. Here, we describe a network of biomarkers that capture both cellular senescence and T cell function and have shown their association with early indicators of physiologic decline. Given a number of recent studies demonstrating that T cells, specifically CD8+ T cells, are critical for immune surveillance of senescent cells (ref), our work provides an early description of markers that can define the relationship between these domains in humans.

Biomarkers in the SapereX network were selected based on previous studies (ref^12^, Justice et al, in print; Sapere Bio, unpublished). Selection criteria included an independent association with p16 and each marker’s capacity to capture distinct as well as potentially overlapping biological functions of the adaptive immune system. Multiple markers of presumably similar function (e.g., T cell exhaustion) were screened for association with p16 as well as the ability to change with longitudinal changes in p16. LAG3 and CD244 were both described as markers of T cell exhaustion and LAG3 is the newest target of immuno-oncology therapies.

CD244 appears to have a distinct role in T cell function, accumulating with age in CD8+ T cells where it is associated with increased proliferation and apoptosis, and terminal differentiation. In addition, CD244 has been shown to be an inhibitor of autophagy by directly interacting with and suppressing activity of the Beclin-1/Vps34 complex^23^. Autophagy is essential for the function of T cells including effector CD8+ T cell survival and memory formation and its activity declines with age^24,25^. While there is a possibility that the immune biomarkers we chose do not fully capture the ability of T cells to surveil senescent cells, our finding that immune biomarkers (LAG3, CD244, CD28) as well as p14 are strongly associated with immune resilience, ie, the ability of the organism to withstand immune challenge and resist inflammation, reinforce the importance of the selected markers. Our data are consistent with a study by Burd et al in healthy adults where p16 expression in T cells was associated with CD28 and CD244 and not other markers of T cell function such as KLRG1, CD57, CD45RA, and the PD-1 pathway^11^.

While SapereX immune markers were independently associated with p16, their relationship is likely to be highly interconnected. For example, in patients with cardiovascular disease, a second-degree interaction between p16 and CD244 was required to predict surgery-associated adverse events^12^. Our study design does not allow us to infer directionality in the relationship between cellular senescence and immune function. While dysfunction in the immune system can lead to accumulation of senescent cells, there is also evidence that accumulation of senescent cells can damage the immune system by inducing T cell exhaustion and reducing T cell proliferation^26,27^. Interestingly, of all the SapereX biomarkers, p14 is most strongly associated with p16, yet clusters with markers of T cell function in the PCA analysis and is associated with immune resilience. It is possible that p14 modulates both domains of the SapereX network-cellular senescence and immune function. p14 may affect cellular senescence through regulation of apoptosis via the p53/p21 axis^28^. Consistent with its putative role in cellular senescence, continuous p14 expression was required for cellular commitment to senescence in the epidermis^29^. Likewise, p14 may regulate immune function. Studies have shown that induction of p14 led to mitochondrial dysfunction and T cell exhaustion in tumor-infiltrating lymphocytes^30^.

Biomarkers in the SapereX network were mostly discordant with other markers of biological aging, including methylation clocks, telomere shortening, and GlycanAge. While global epigenetic changes are associated with the development and maintenance of the senescent phenotype^31–33^, the most commonly used methylation clocks do not correlate with cellular senescence (Sedrak 2023, in print, and Fig 3) or SapereX immune biomarkers. Telomere shortening was modestly associated with CD244 (r=0.34), possibly due to the link between CD244 and apoptosis. Not surprisingly, telomere shortening was also associated with p16 (r=0.28), but not other markers. Interestingly, neither GlycanAge nor the intrinsic epigenetic clock (TruDiagnostic), putative biomarkers of immune function and chronic inflammation, was associated with immune resilience (Figure 3). Likewise, neither GlycanAge nor the intrinsic epigenetic clock was associated with components of the SapereX network.

With increasing attention on interventions that may extend human healthspan and ongoing clinical trials to alter organismal cellular senescence and other aging pathways, it is increasingly important to understand the dynamics of these processes in humans. A current hypothesis in the field is that a decline in immune function may precede accumulation of senescent cells (ref). In some patients, addressing immune dysfunction may provide an indirect way of targeting cellular senescence and improving aging trajectory. Alternatively, understanding the interplay between the immune system and cellular senescence may improve patient selection and avoid harm in clinical trials. For example, senolytic therapy may be inappropriate for patients with already low cellular senescence and normal immune biomarkers, while patients with low cellular senescence but dysregulated immune function may benefit from interventions targeting the immune system. And vice versa, patients with high cellular senescence but normal immune biomarkers may be better suited to a senolytic approach absent an immune intervention.

Our study has some limitations including a relatively small sample size, particularly for clinical variables collected for a subset of participants, e.g insulin resistance. Another limitation is the cross-sectional design. Longitudinal data are needed to validate the presence of a temporal relationship between biomarkers of immune function and cellular senescence and that work is ongoing. In addition, CD11b+ dendritic cells appear to be essential mediators of surveillance of senescent cells by CD8+ T cells^34^. In this study, we have not captured measures of dendritic cell activation or function. However, we have previously demonstrated that plasma levels of suPAR, a protein secreted mostly by the immune cells^35^, contributes to the predictive ability of SapereX to identify clinical vulnerabilities mediated by uncontrolled inflammation in the setting of cardiac surgery.

There is a time-honored adage in medicine - ‘what gets measured, gets managed’. Aging, in its foundational stages, is rarely managed in part because clinically relevant molecular measures are lacking^36^. This despite growing recognition that progressive and dramatic loss of physiological resilience precedes frailty and chronic disease by many decades. The ability to measure and map changes in immune aging through cellular senescence and T cell function creates the potential for improved safety, efficacy, and personalization of diverse interventions aimed at improving health span, longevity, and functional performance throughout life.

## DATA AVAILABILITY

Due to the informed consent and data privacy policies, the clinical data are not publicly available, i.e., accessible for anyone, for any purpose without a review by the Central IRB on a project-by-project basis.

